# The Chronic Kidney Disease and Acute Kidney Injury Involvement in COVID-19 Pandemic: A Systematic Review and Meta-analysis

**DOI:** 10.1101/2020.04.28.20083113

**Authors:** Ya-Fei Liu, Zhe Zhang, Xiao-Li Pan, Guo-Lan Xing, Ying Zhang, Zhang-Suo Liu, Sheng-Hao Tu

## Abstract

**Aim:** The aim of this study was to uncover whether kidney diseases were involved in COVID-19 pandemic from a systematic review.

**Methods:** The studies reported the kidney outcomes in different severity of COVID-19 were included in this study. Standardized mean differences or odds ratios were calculated by employing Review Manager meta-analysis software.

**Results:** Thirty-six trials were included in this systematic review with a total of 6395 COVID-19 patients. The overall effects indicated that the comorbidity of chronic kidney disease (CKD) (OR = 3.28), complication of acute kidney injury (AKI) (OR = 11.02), serum creatinine (SMD = 0.68), abnormal serum creatinine (OR = 4.86), blood urea nitrogen (SMD = 1.95), abnormal blood urea nitrogen (OR = 6.53), received continuous renal replacement therapy (CRRT) (OR = 23.63) was significantly increased in severe group than that in nonsevere group. Additionally, the complication of AKI (OR = 13.92) and blood urea nitrogen (SMD = 1.18) were remarkably elevated in critical group than that in severe group.

**Conclusion:** CKD and AKI are susceptible to occur in patients with severe COVID-19. CRRT is applied frequently in severe COVID-19 patients than that in nonsevere COVID-19 patients. The risk of AKI is higher in critical group than that in severe group.

## 1. Introduction

In December, 2019, 41 hospitalized patients had been identified as having laboratory-confirmed 2019-novel coronavirus (2019-nCoV)-related infection in Wuhan, Hubei, China^[1]^. The 2019-nCoV was discovered by whole-genome sequencing, direct PCR, and culture^[2]^. The next-generation sequencing of 2019-nCoV confirmed that it is sufficiently divergent from severe acute respiratory syndrome (SARS)-CoV and considered as a novel human-infecting betacoronavirus^[3]^. Moreover, structural analysis revealed that 2019-nCoV might bind to the angiotensin converting enzyme 2 (ACE2) receptor in humans^[3]^. The 2019-nCoV causing the current outbreak of coronavirus disease has been named SARS-CoV-2 by the International Committee Taxonomy of Viruses. On 30 January 2020, the World Health Organization (WHO) Director-General declared the 2019-nCoV outbreak (https://www.who.int). On 11 March 2020, the WHO declared novel coronavirus disease (COVID-19) to be a global pandemic (https://www.who.int). The SARS-CoV-2 promptly spread across China and around the world. As of April 14, 2020, a total of more than 1,848,000 laboratory-confirmed cases had been documented worldwide. The most common clinical characteristics were fever (43.8% on admission and 88.7% during hospitalization) and cough (67.8%)^[4]^. The single-cell RNA sequencing datasets indicated that ACE2 was mainly expressed in lung type II alveolar cells, colon colonocytes, ileum ECs, and proximal tubule cells^[5,6]^. In addition, SARS-CoV-2 was detected in urine, blood, anal swab, and oropharyngeal swab from nine patients with COVID-19 who were retested by qRT-PCR^[7]^. The above evidence revealed that SARS-CoV-2 can affect respiratory system, digestive system, hematological system, and urinary system which were associated with the clinical features of COVID-19. Moreover, the single-cell transcriptome analysis identified that ACE2 and transmembrane protease serine 2 (TMPRSS2) genes are relatively high coexpression in podocytes and proximal tubule cells^[8]^.

A prospective cohort study of 701 patients with COVID-19 showed that the prevalence of increased serum creatinine, elevated blood urea nitrogen, and estimated glomerular filtration (eGFR) < 60 ml/min/1.73m^2^ were 14.4, 13.1 and 13.1%, respectively^[9]^. Meanwhile, acute kidney injury (AKI) occurred in 5.1% COVID-19 patients^[9]^. A meta-analysis including 1389 COVID-19 patients indicated that chronic kidney disease (CKD) was consistent with severe COVID-19^[10]^. Meanwhile, the China professionals point out that AKI cannot be ignored in the diagnosis and treatment of COVID-19^[11]^. However, the study including 116 COVID-19-confirmed patients demonstrated that SARS-CoV-2 infection does not result in AKI, or aggravate CKD in the COVID-19 patients^[12]^. Therefore, it is stringent to perform a systematic review to verify whether the kidney diseases are involved in COVID-19.

## 2. Materials and methods

The Preferred Reporting Items for Systematic Review and Meta-Analyses (PRISMA) statement was employed to design and report the study^[13]^.

### 2.1. Search Strategy. The following English databases were retrieved to confirm trials

PubMed and medRxiv.gov. In addition, the Chinese databases, such as the CNKI Database and WanFang Database were searched. All of the databases were searched from their available dates of inception to the latest issue (April 13, 2020).

Different search strategies were integrated as follows. For the English databases, free text terms were employed, such as “SARS-CoV-2” or “2019-nCoV” or “COVID-19” or “novel coronavirus pneumonia” and “kidney” or “renal”. For the Chinese databases, free text terms were applied for “xin xing guan zhuang bing du” (which means SARS-CoV-2 in Chinese) or “xin xing guan Zhuang bing du fei yan” (which means COVID-19 in Chinese). The reference lists of relevant publications were also searched to identify extra studies.

### 2.2. Selection Criteria

Inclusion criteria: (1) study type: case series study, cohort study, or prospective study which reported the connection between kidney involvement and COVID-19 irrespective of publication status or language; (2) subjects: adult patients diagnosed with COVID-19; (3) outcomes: COVID-19 patients with CKD comorbidity, AKI, serum creatinine, blood urea nitrogen, or received continuous renal replacement therapy (CRRT).

The case reports, reviews, meta-analysis, or studies regarding renal replacement therapy regularly were excluded. For repeat studies whose preprints were published in medRxiv.gov, the final version studies were included. The literatures were selected by two reviewers (YF Liu and Z Zhang) independently. The flowchart of the study selection has been drafted in accordance to the PRISMA principle.

### 2.3. Data Extraction and Management

The data were extracted by two independent reviewers (XL Pan and GL Xing), and contradictions were resolved by consensus or were judged by another author. The data presented as median and interquartile range was transformed into mean and standard deviation according to the formula below (http://www.math.hkbu.edu.hk/~tongt/papers/median2mean.html).

The studies’ quality was evaluated according to the Newcastle-Ottawa scale (NOS) which assess the quality of nonrandomised studies in meta-analyses^[14]^. The quality of the included studies was estimated independently by two investigators (SH Tu and Y Zhang). The NOS scores ≥ 6 were considered high quality studies. The authors were connected to clarify vagueness or absence of the data, and related data were extracted by consensus if the authors were unavailable.

### 2.4. Data Synthesis and Analysis

The Review Manager meta-analysis software, version 5.3 was applied to analyze the indicators. The standardized mean differences (SMDs) and 95% confidence intervals (CIs) were calculated for continuous data. The odds ratios (ORs) and 95% CIs were calculated for dichotomous data. Heterogeneity was estimated through the chi-square test and Higgins I^2^ test. A fixed-effect model was applied if the studies were sufficiently alike (*P* > 0.10); otherwise, a random-effects model was employed. A Z score was calculated to detect the overall effect, with significance set at *P* < 0.05. Publication bias was evaluated by funnel plot if the number of included studies > 10.

Two subgroup analyses were performed to diminish the clinical heterogeneity according to the disease severity: severe and nonsevere groups, critical and severe groups.

## 3 Results

### 3.1. Study Selection

The process of study selection was showed in Fig. 1. After filtering, COVID-19 patients were divided into severe and nonsevere groups in 28 studies^[1,4,15–40]^. In 8 studies, the COVID-19 patients were classified as more than two groups according to the disease severity which included severe and critical COVID-19 patients^[41–48]^. Finally, 36 studies were included in the study. The characteristics of the studies were indicated in Table 1. Together, 6395 COVID-19 patients were included in the study.

**Fig. 1.**
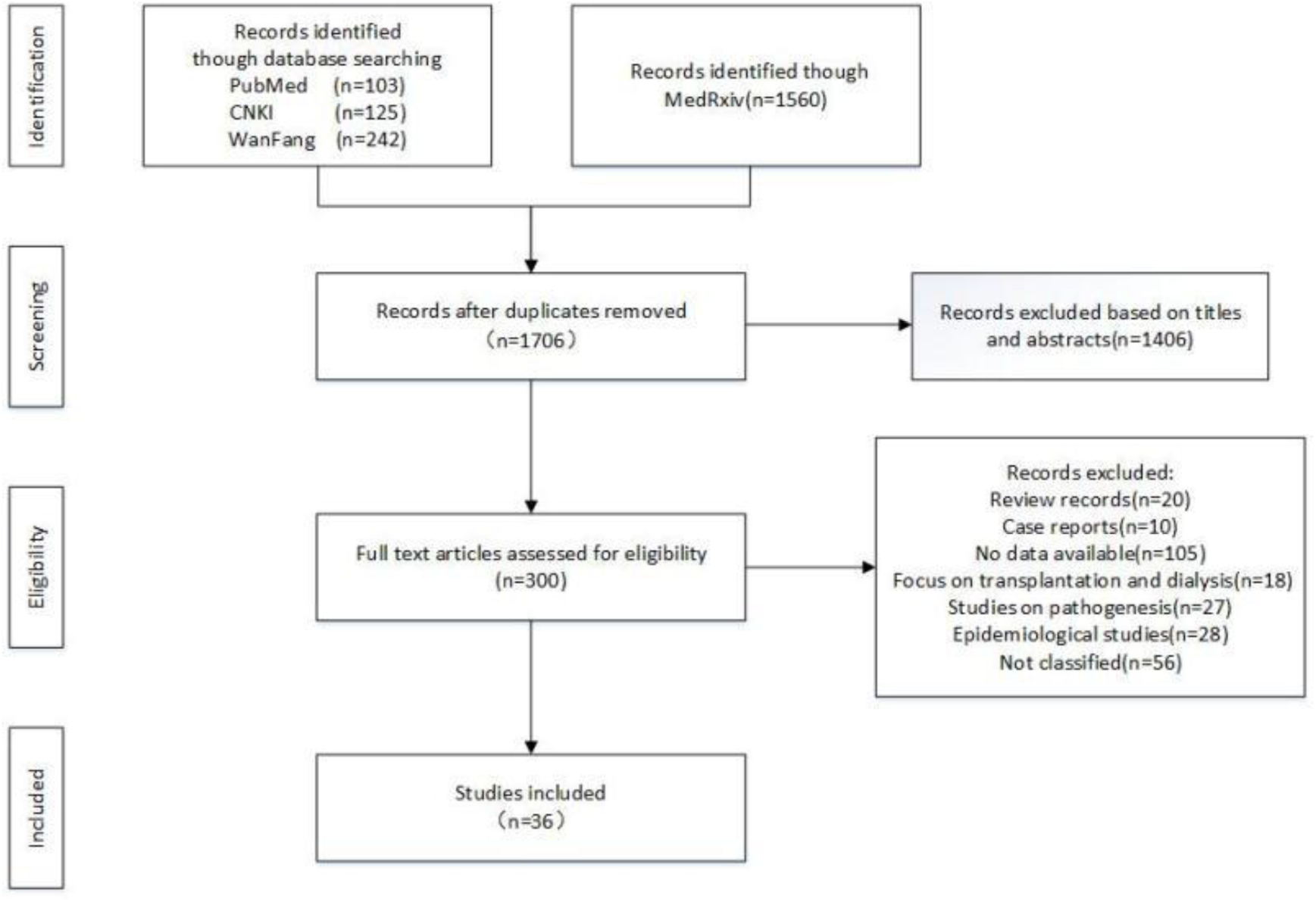
Flowchart of study selection.

**Table 1:**
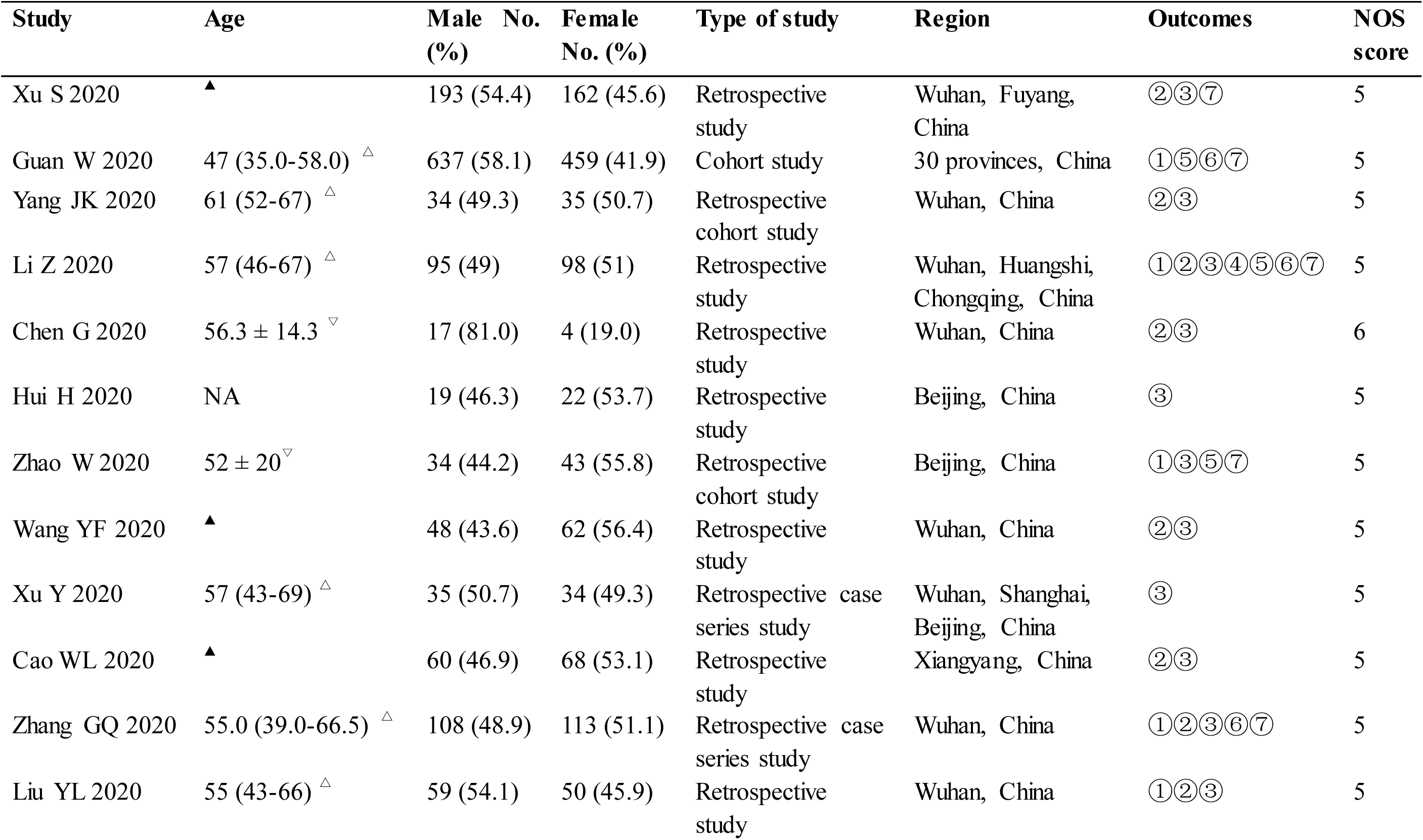
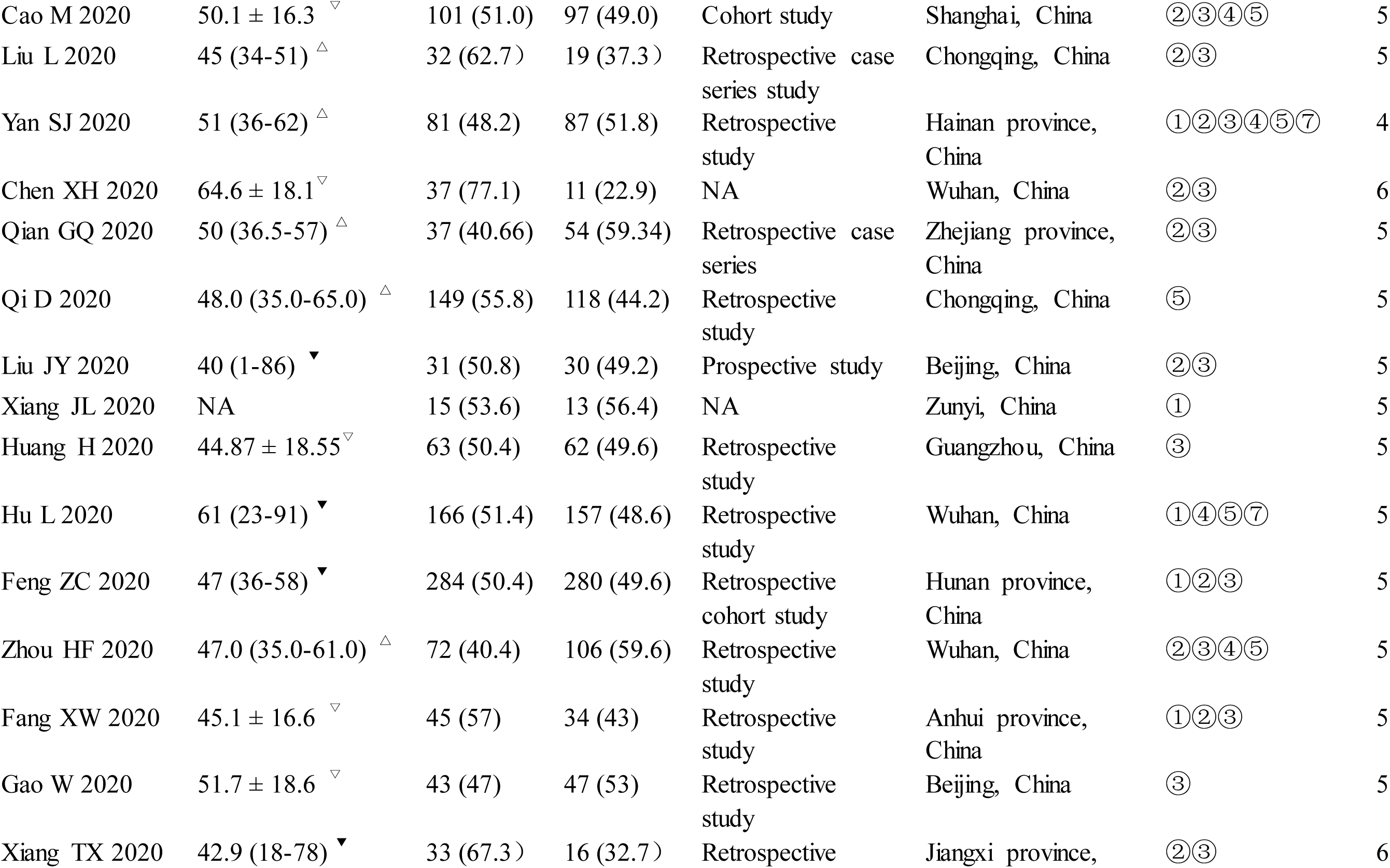
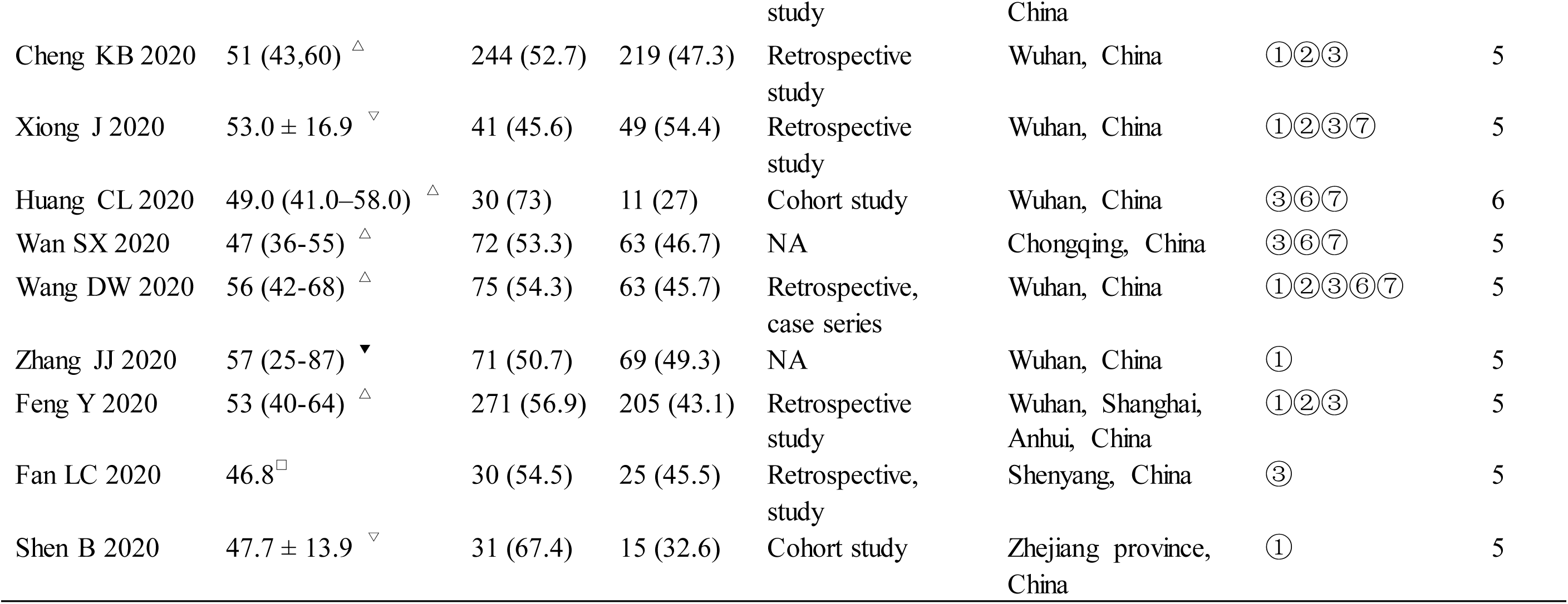
Clinical and demographic characteristics of the patients with COVID-19. Note: Age was expressed as different levels▴ or median and interquartile range▵ or median and range▾ or mean ± standard deviation▿ or mean▢. NA: not available. ① Chronic kidney disease; ② Blood urea nitrogen; ③ Serum creatinine; ④ Abnormal blood urea nitrogen; ⑤ Abnormal serum creatinine; ⑥ Continuous renal replacement therapy; ⑦ Acute kidney injury;

### 3.2. Study Descriptions

All the included studies were performed in China. Five studies were published in Chinese^[30–32,47,48]^, and the others were published in English. Eight studies were conducted as multi-center trials^[4,15,19,23–25,29,45]^. The patients with COVID-19 divided into ICU care and no ICU care in four studies were included in the severe and nonsevere groups^[1,36,37,39]^. The COVID-19 patients divided into ARDS (acute respiratory distress syndrome) and non-ARDS in one study were also included in the severe and nonsevere groups^[38]^. Two studies combined the data of severe and critical patients together, and they were also included in subgroup severe and nonsevere groups^[30,31]^.

### 3.3. Quality of the Included Studies

As indicated in Table 1, the majority of included studies were poor quality except four studies whose NOS scores are 6^[1,16,31,43]^. The NOS scores of other 31 trials are 5, and only one trial’s NOS scores were 4.

### 3.4. Publication Bias

As the included studies of three outcomes > 10 in severe and nonsevere groups, the funnel plots were conducted to estimate the publication bias (Fig. 2). The funnel plot of CKD was symmetrical, suggesting that there was no publication bias. However, the funnel plot of blood urea nitrogen was a little asymmetrical, implying that the publication bias existed to some extent.

**Fig. 2.**
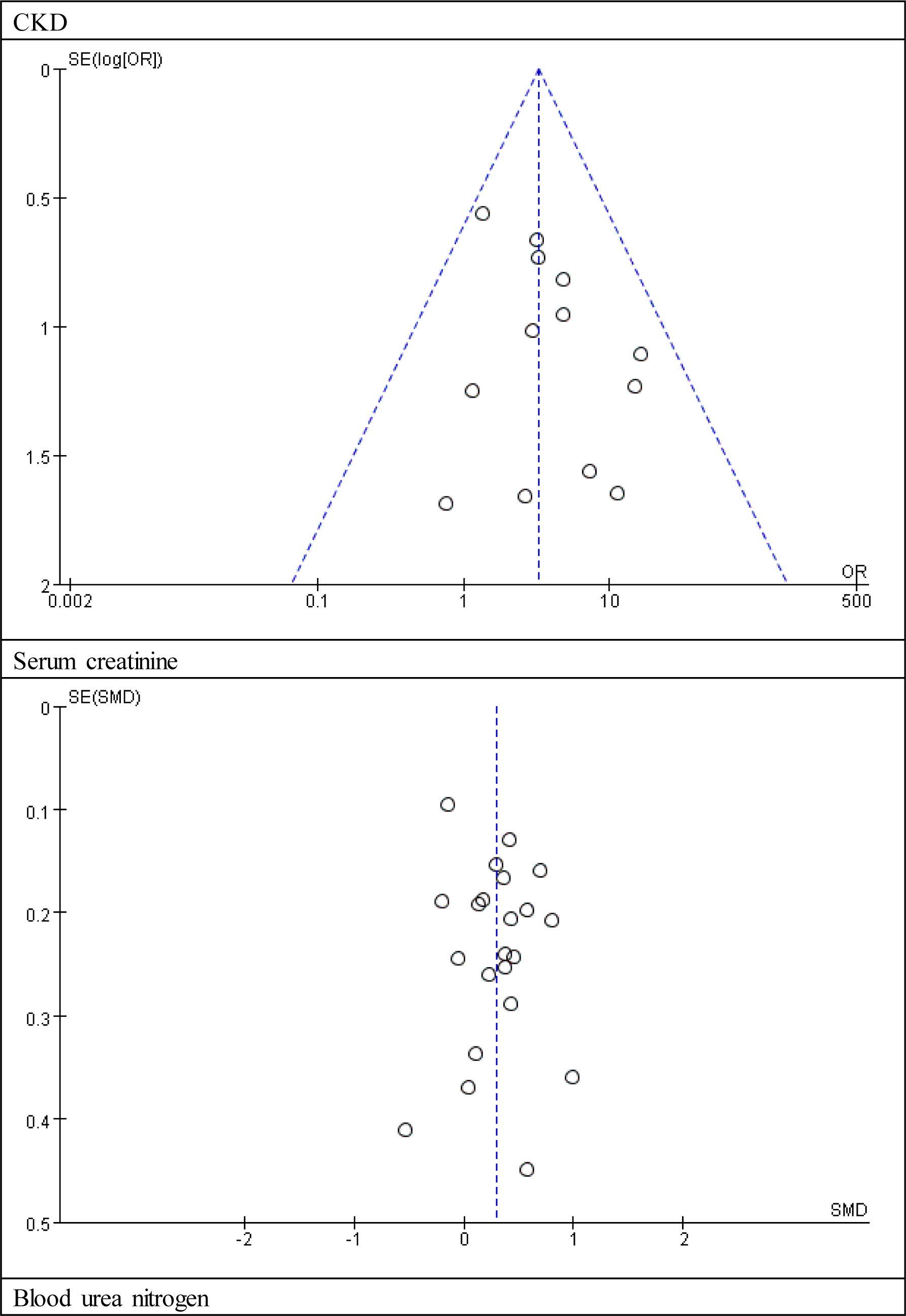
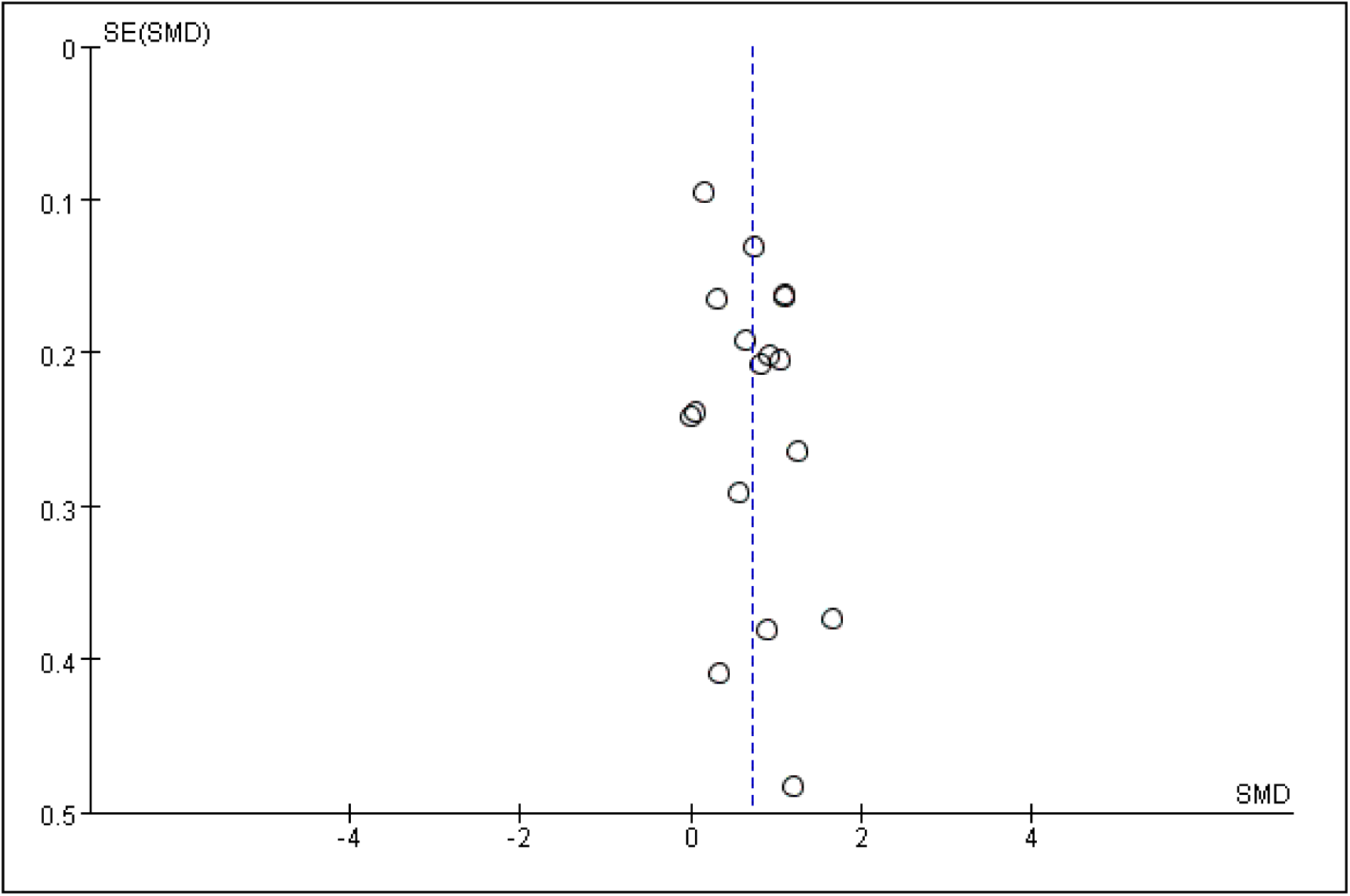
The funnel plots. Note: CKD: chronic kidney disease.

### 3.5. Effects of Outcomes

#### 3.5.1. Severe and nonsevere groups

##### 3.5.1.1 The outcome of comorbidity of CKD

As illustrated in Fig. 3, the number of participants ranged from 21 to 1099. There was no statistical heterogeneity between the studies (*P* = 0.72). Thirteen studies mentioned comorbidity of CKD (including 3325 subjects), and pooled results indicated that comorbidity of CKD was significantly increased in severe group than that in nonsevere group (*P* < 0.00001, OR = 3.28, 95% CI: 2.0 to 5.37).

**Fig. 3.**
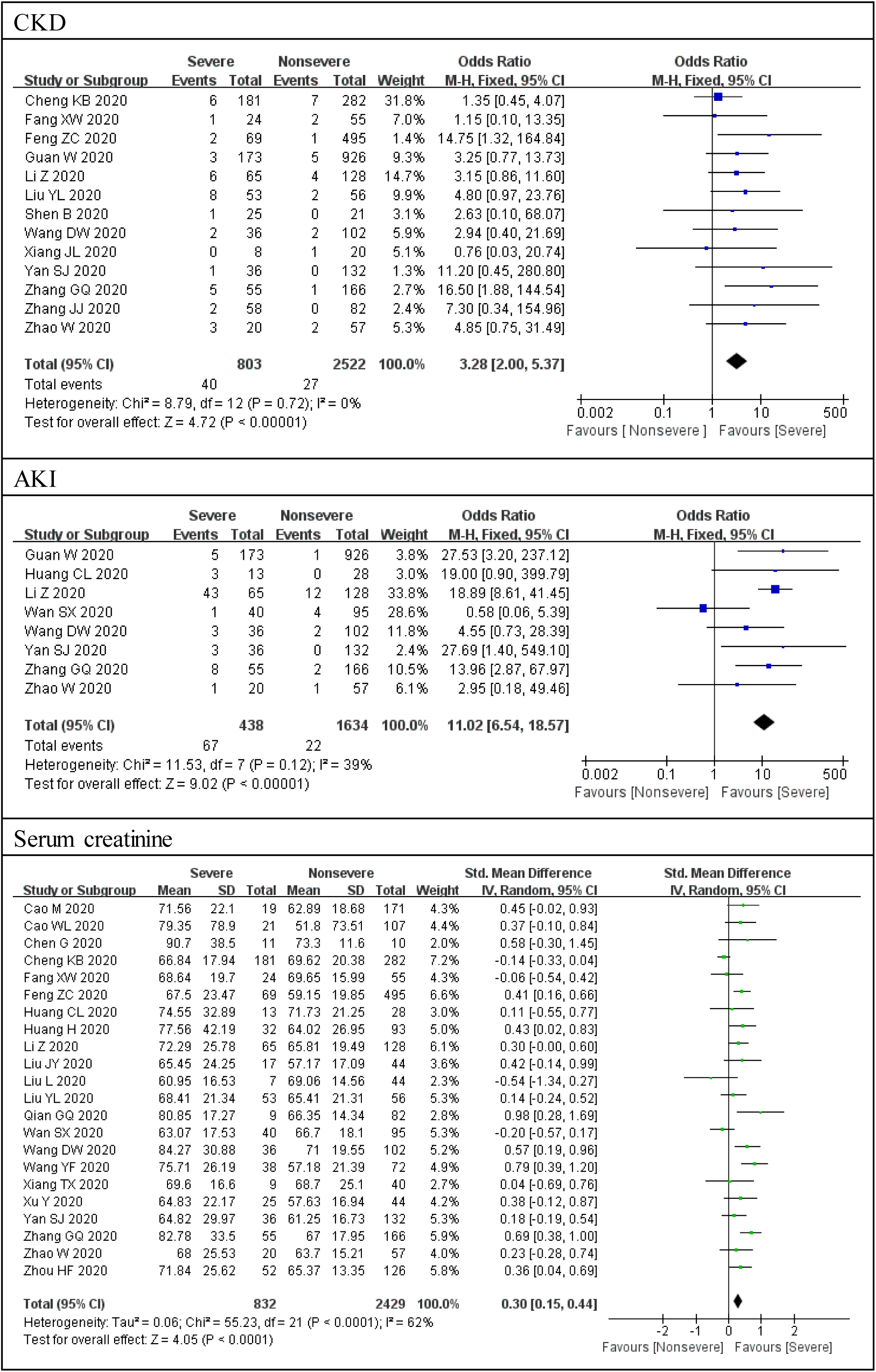
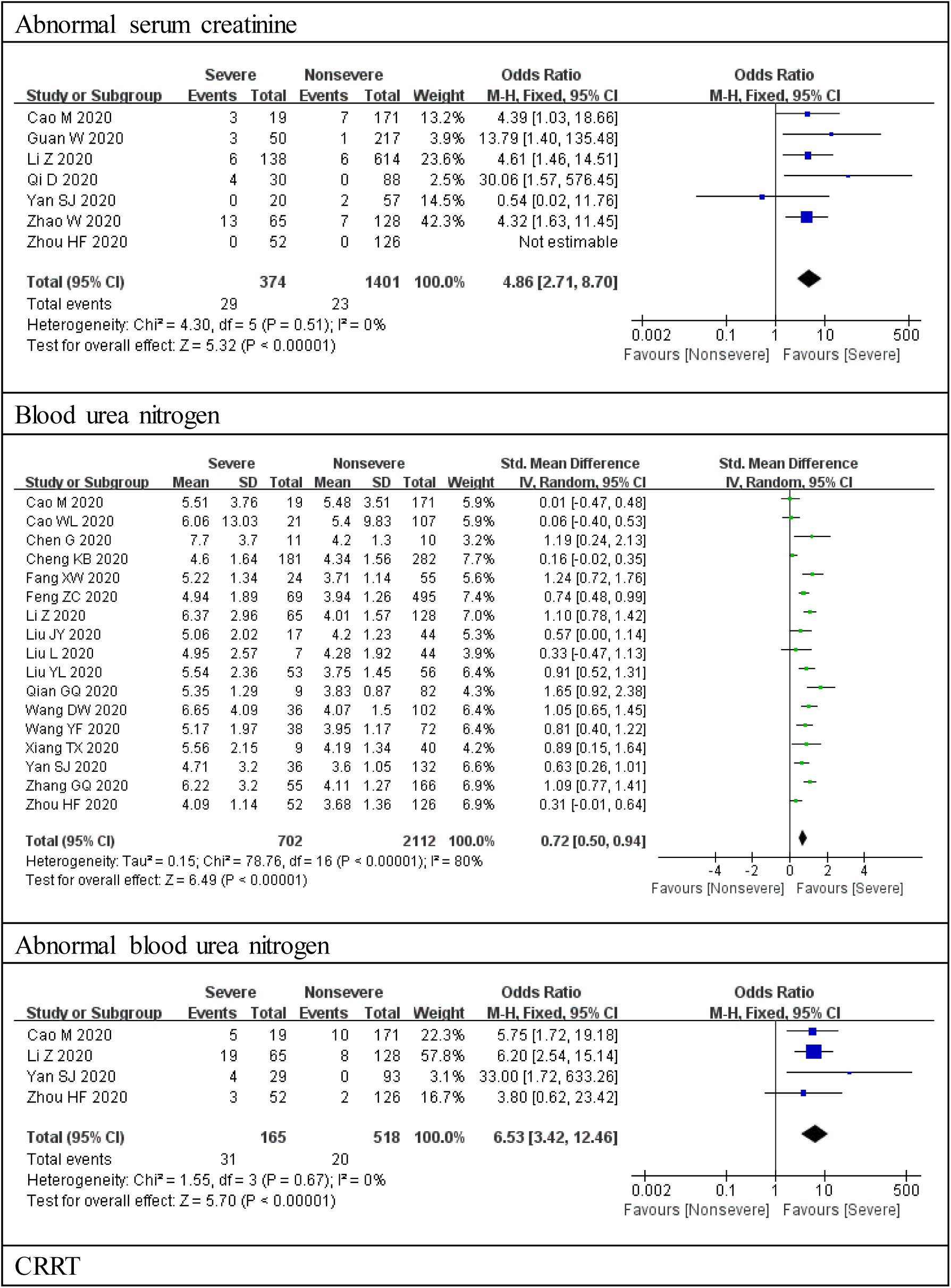
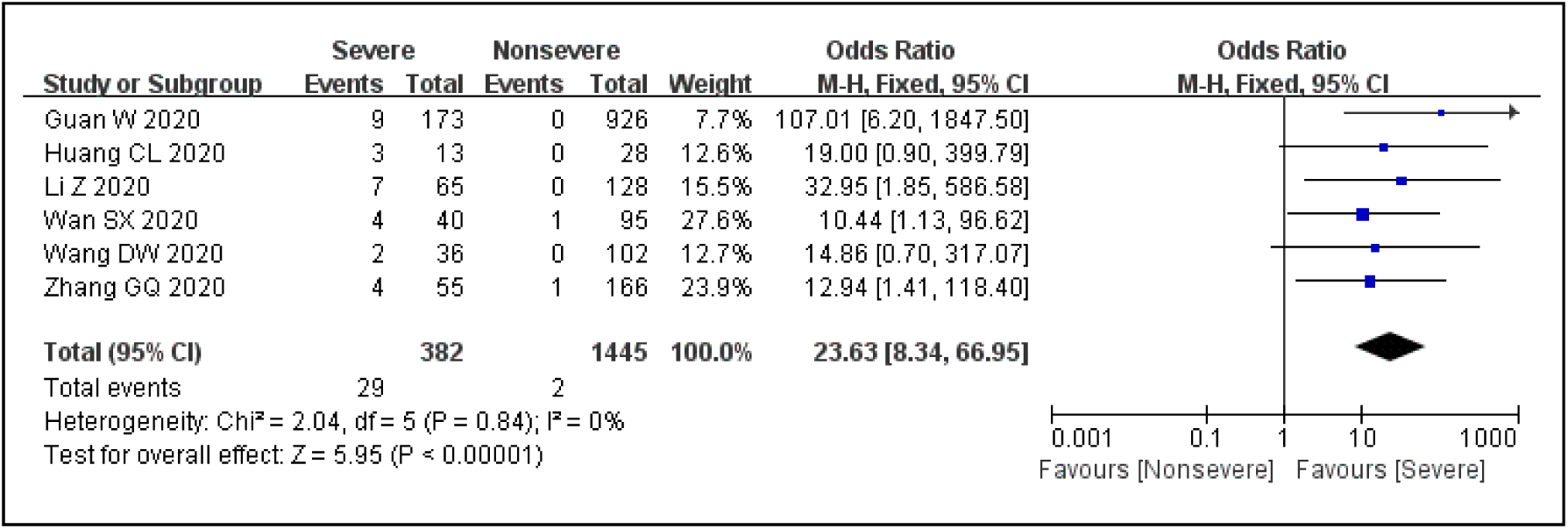
Forest plots for the severe and nonsevere subgroup. Note: CKD: chronic kidney disease; AKI: acute kidney injury; CRRT: continuous renal replacement therapy.

##### 3.5.1.2. The outcome of complication of AKI

As showed in Fig. 3, the number of participants ranged from 41 to 1099. There was no statistical heterogeneity between the studies (*P* = 0.12). Eight studies reported complication of AKI (involving 2072 subjects), and complication of AKI was significantly elevated in severe group than that in nonsevere group (*P* < 0.00001, OR = 11.02, 95% CI: 6.54 to 18.57).

##### 3.5.1.3. The outcome of serum creatinine

As illustrated in Fig. 3, the number of participants ranged from 21 to 564. There was statistical heterogeneity between the studies (*P* < 0.00001). Twenty-two studies measured serum creatinine (including 3261 subjects), and pooled results showed that serum creatinine was remarkably increased in severe group than that in nonsevere group (*P* = 0.0005, SMD = 0.68, 95% CI: 0.3 to 1.06).

##### 3.5.1.4. The outcome of abnormal serum creatinine

As indicated in Fig. 3, the number of participants ranged from 77 to 752. There was no statistical heterogeneity between the studies (*P* = 0.51). Seven studies documented abnormal serum creatinine (including 1775 subjects), and severe group had a higher ratio of abnormal serum creatinine than nonsevere group (*P* < 0.00001, OR = 4.86, 95% CI: 2.71 to 8.7).

##### 3.5.1.5. The outcome of blood urea nitrogen

As illustrated in Fig. 3, the number of participants ranged from 21 to 564. There was statistical heterogeneity between the studies (*P* < 0.00001). Seventeen studies assayed blood urea nitrogen (including 2814 subjects), and pooled results revealed that blood urea nitrogen was significantly elevated in severe group than that in nonsevere group (*P* < 0.00001, SMD = 1.95, 95% CI: 1.23 to 2.66).

##### 3.5.1.6. The outcome of abnormal blood urea nitrogen

As showed in Fig. 3, the number of participants ranged from 122 to 193. There was no statistical heterogeneity between the studies (*P* = 0.67). Four studies documented abnormal blood urea nitrogen (involving 683 subjects), and severe group had a higher ratio of abnormal blood urea nitrogen than nonsevere group (*P* < 0.00001, OR = 6.53, 95% CI: 3.42 to 12.46).

##### 3.5.1.7. The outcome of receiving CRRT

As illustrated in Fig. 3, the number of participants ranged from 41 to 1099. There was no statistical heterogeneity between the studies (*P* = 0.84). Six studies reported receiving CRRT (including 1827 subjects), and severe group had higher ratio of receiving CRRT than nonsevere group (*P* < 0.00001, OR = 23.63, 95% CI: 8.34 to 66.95).

#### 3.5.2. Critical and Severe groups

##### 3.5.2.1 The outcome of comorbidity of CKD

As illustrated in Fig. 4, the number of participants ranged from 50 to 166. There was no statistical heterogeneity between the studies (*P* = 0.88). Three studies reported comorbidity of CKD (including 346 subjects), and pooled results indicated that there was no significant difference between the two groups (*P* = 0.77, OR = 1.3, 95% CI: 0.23 to 7.16).

**Fig. 4.**
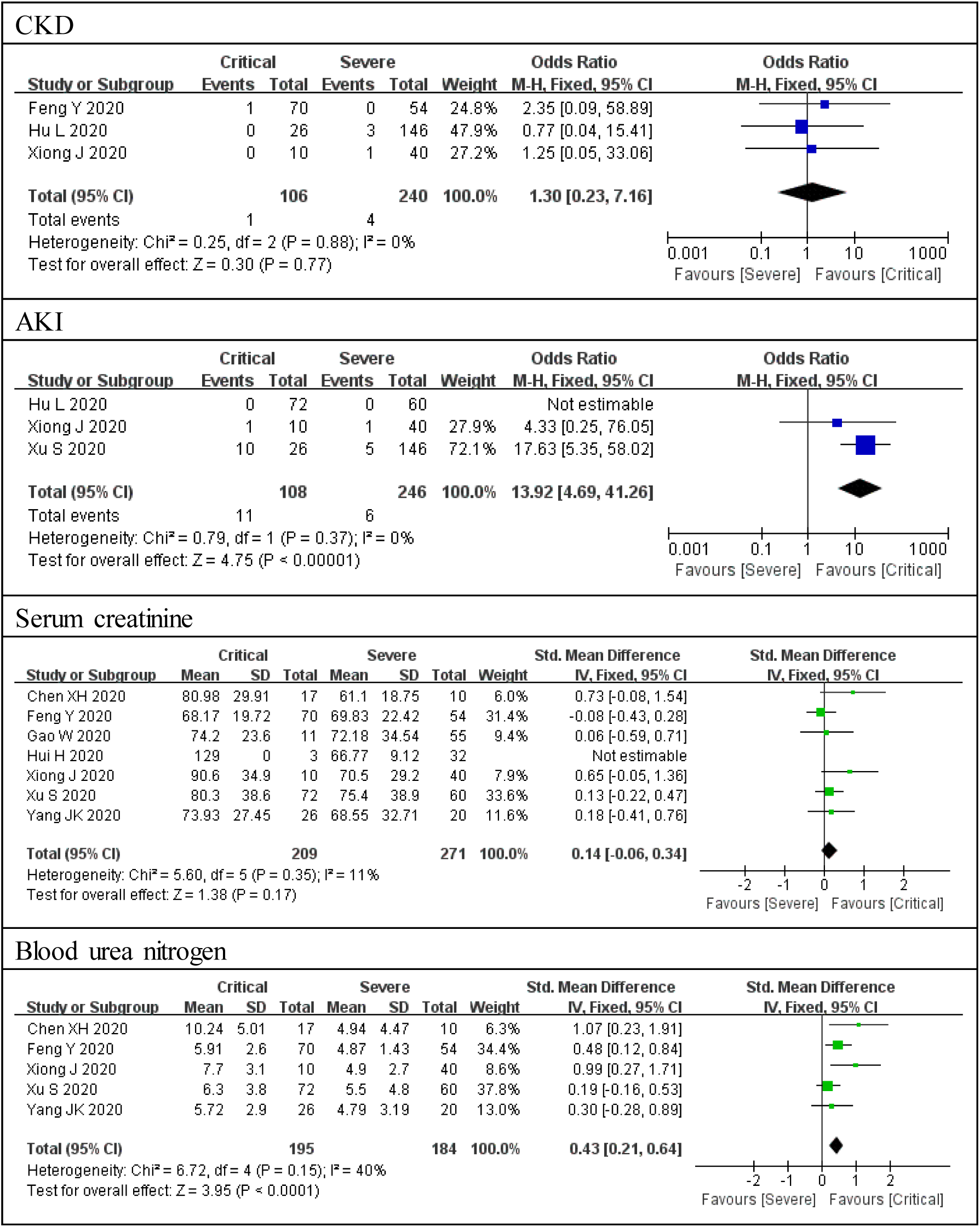
Forest plots for the critical and severe subgroup. Note: CKD: chronic kidney disease; AKI: acute kidney injury.

##### 3.5.2.2. The outcome of complication of AKI

As indicated in Fig. 4, the number of participants ranged from 50 to 172. There was no statistical heterogeneity between the studies (*P* = 0.37). Three studies documented complication of AKI (involving 354 subjects), and the complication of AKI was significantly elevated in critical group than that in severe group (*P* < 0.00001, OR = 13.92, 95% CI: 4.69 to 41.26).

##### 3.5.2.3. The outcome of serum creatinine

As illustrated in Fig. 4, the number of participants ranged from 27 to 132. There was statistical heterogeneity between the studies (*P* < 0.00001). Seven studies determined serum creatinine (including 480 subjects), and pooled results indicated that there was no significant difference between the two groups (*P* = 0.13, SMD = 0.43, 95% CI: −0.13 to 0.99).

##### 3.5.2.4. The outcome of blood urea nitrogen

As showed in Fig. 4, the number of participants ranged from 27 to 132. There was statistical heterogeneity between the studies (*P* < 0.00001). Five studies measured blood urea nitrogen (including 379 subjects), and pooled results showed that blood urea nitrogen is strikingly increased in critical group than that in severe group (*P* = 0.002, SMD = 1.18, 95% CI: 0.44 to 1.91).

Only one study reported abnormal blood urea nitrogen and abnormal serum creatinine, so the result was not pooled. CRRT was not documented in this subgroup.

## 4. Discussion

To our knowledge, this is the first research which comprehensively illustrates the kidney involvement in COVID-19 pandemic. This systematic review observed that the CKD and AKI were involved in the COVID-19. Compared with nonsevere group, the comorbidity of CKD (OR = 3.28) and complication of AKI (OR = 11.02) were significantly elevated in severe group. Meanwhile, AKI occurred in 4.3% COVID-19 patients, and 21.1% were severe COVID-19 patients. The complication of AKI is lower than 5.1% which was reported in a previous cohort study, of which 42.7% were severely ill^[9]^. The different disease severity and sample size may contribute to the difference. The comorbidity of CKD is similar to previous meta-analysis (including 1389 participants) which showed that CKD was significantly associated with severe COVID-19 (OR = 3.03), among which 19.7% were defined as severe COVID-19^[10]^.

Furthermore, the serum creatinine, ratio of abnormal serum creatinine, blood urea nitrogen, ratio of abnormal blood urea nitrogen were remarkably elevated in severe group than that in nonsevere group, with SMD = 0.68, OR = 4.86, SMD = 1.95, OR = 6.53, respectively. The OR regarding abnormal serum creatinine and abnormal blood urea nitrogen were lower than that in the severe and nonsevere groups with regard to AKI. The AKI also occurred in normal serum creatinine and blood urea nitrogen, which could lead to the higher OR of AKI.

The CRRT is recommended in severe or critical patients because it reduces cytokine damage, removes endotoxin, and improves cardiac function and kidney function^[49]^. Hence, the severe group had higher ratio of receiving CRRT than nonsevere group (OR = 23.63).

The critical group had a higher ratio of AKI than that in severe group (OR = 13.92) which is higher than subgroup severe and nonsevere groups (OR = 11.02). Furthermore, the blood urea nitrogen was increased in critical group than that in severe group (SMD = 1.18). However, there is no significant difference between the two groups with regard to comorbidity of CKD. Owing to immunodepression of CKD, cytokine storm is weakened and CKD patients are not vulnerable to critical COVID-19, which could explain the result. Meanwhile, there was no significant increase in serum creatinine in critical group. We speculated that the CRRT was frequently applied in critical group, so the serum creatinine was not notably elevated in critical group. In addition, AKI is diagnosed not only by abnormal serum creatinine but also the change of serum creatinine and urine volume.

There are lots of reasons which give rise to the kidney impairment in COVID-19. First, SARS-CoV-2 viral load induce cytotoxicity of renal resident cells. The endothelial cell swelling, renal tubular epithelial cell swelling, vacuolar degeneration, and drop were observed with light microscope of renal biopsy in three COVID-19 cases^[50]^. Second, the fever, vomit, diarrhea, and shock are likely to cause kidney hypoperfusion. Third, Serum SARS-CoV-2 viral load (RNAaemia) is strongly associated with cytokine storm^[43]^. cytokine storm participated in SARS-CoV-2 infection^[51]^. A renal transplanted patient with COVID-19 showed a mild disease because immunosuppression of renal transplantation can “be protective”^[52]^. Fourth, the nephrotoxicity of some drugs, such as non-steroidal anti-inflammatory drugs, antivirals, antibiotics, and antifungal could lead to kidney damage. Fifth, organ crosstalk such as cardiorenal syndrome, hypoxia, and rhabdomyolysis also could cause kidney impairment^[49]^. Sixth, mechanical ventilation is able to result in AKI^[53]^. Seventh, the elderly patients with comorbidities such as hypertension, diabetes, chronic cardiology disease, or chronic liver disease are susceptible to trigger secondary kidney diseases which may not be confirmed before admission.

Several limitations of the included studies should be concerned. First, all the included studies did not stage CKD. Different CKD stage could have different risks for COVID-19 and have different prognosis. Second, only one study^[9]^ reported the AKI stage which reflects the AKI severity, progression, and prognosis. AKI occurred primarily or AKI on CKD is unknown for the included studies. Third, all the included studies did not qualify the 24h total urine protein. Therefore, it is difficult to judge the severity of potential kidney diseases. Fourth, all the included studies did not measure the 24h urine volume or the change of urine volume which is beneficial for the diagnosis of AKI. Only two studies reported the urine routine^[15,39]^. Finally, the outcomes of AKI were unclear.

Several limitations of this study should be noticed. First, all the included trials were conducted in Chinese populations, which inferred a high risk of selection bias. Second, most of the studies were of poor quality except four studies. Third, the heterogeneity was significant in terms of serum creatinine and blood urea nitrogen. The different disease severity, different sample size, different detection methods, and reference range were responsible for the heterogeneity. Fourth, the children or teenagers COVID-19 patients were probably included in this study because the included studies did not clearly restrict to adult COVID-19 patients. Fifth, the quality of the most of included studies was low which may weaken the evidence level. Finally, the most important criteria eGFR was reported in only three studies^[30,37,47]^. In view of this, we should carefully interpret all of the conclusions due to the considerable poor quality and clinical variety of the studies.

A series of recommendations are given: first, detailed kidney-specific examinations are necessary for the COVID-19 patients with proteinuria or AKI, such as urine routine, 24h total urine protein, urine Bence-Jones protein, immunoglobulins, complements, antinuclear antibody, anti-extractable nuclear antigen antibodies, antineutrophil cytoplasmic antibodies, anti-glomerular basement membrane antibodies, anti-phospholipase A2 receptor antibodies, estimated glomerular filtration, ultrasound of urinary system. Second, recent studies focused on the tubular epithelial cell injury, renal biopsy and renal pathology including light microscope, electron microscope, and immunofluorescence are essential to determine whether other kidney resident cells are involved in COVID-19. Third, the patients with proteinuria or AKI are needed to follow-up.

## 5. Conclusion

In conclusion, this systematic review suggests that CKD and AKI are involved in severe COVID-19. Severe COVID-19 is susceptible to occur in CKD patients than nonsevere COVID-19. With the severity of COVID-19 increasing, the risk of AKI elevated strikingly. CRRT is applied more frequently in severe group than that in nonsevere group. Clinicians should be cautious of kidney diseases in patients with COVID-19, especially the severe and critical COVID-19. More attention should be paid to monitoring kidney function to avoid end stage renal disease and decease in COVID-19 pandemic.

## Data Availability

The data used to support the findings of this study are available from the corresponding author upon request.

## Funding

This work was funded by the National Natural Science Foundation of China (81701601).

## Declaration of competing interest

All authors state that there is no conflict of interest.

## Acknowledgments

We would like to acknowledgement the National Natural Science Foundation of China for its financial support.

## Notes

### Competing Interest Statement

The authors have declared no competing interest.

